# Children and adolescents with anterior knee pain can show construct validity and reliability with anterior knee pain scale, lower extremity functional scale and visual analogue scale

**DOI:** 10.1101/2025.07.20.25331597

**Authors:** Sarah Hofmeister, Linda Camilleri, Stacey Llewellyn

## Abstract

**Background:** To investigate validity and reliability of the Anterior Knee Pain Scale, Lower Extremity Functional Scale and the Visual Analogue Scale Usual and Worst in 8-16-year-old children and adolescents with symptoms of ‘Anterior knee pain’ commonly present to emergency and orthopaedic outpatients departments at Australian public tertiary/quaternary hospitals

**Method:** 50 participants aged 8-16 years with anterior knee pain participated in this prospective pre- and post-test methodological study. Measures employed included the Anterior Knee Pain Scale (AKPS), the Lower Extremity Functional Scale (LEFS), Visual Analogue Scale Usual (VAS-U) and Visual Analogue Scale Worst (VAS-W). Correlations between these outcome measures and the participant-perceived Global Rating of Change (GRC) categories (incomplete improvement, complete improvement) were determined.

**Results:** *Test-retest reliability:* Intraclass Coefficient (ICC) values indicated good to excellent reliability in VAS-U (0.86), VAS-W (0.70), AKPS (0.98) and LEFS (0.98).

*Validity:* All measures showed convergent validity in comparison to the combined GRC assessment of complete score. The adjusted mean change scores had moderate evidence of a difference in AKPS (p=0.015) and LEFS (0.050); and very strong evidence for difference in VAS-U (p=0.006) and VAS-W (p<0.001) in the improvement groups, with a larger mean change exhibited in the complete improvement group.

*Responsiveness:* ANCOVA results indicated all the outcomes had larger adjusted mean scores in the complete group compared to the incomplete improvement group, suggesting these measures are responsive to measure change.

**Conclusion:** Anterior Knee pain is a common knee condition from child to adults. Just like in adults, questionnaires AKPS, LEFS, VAS-U, and VAS-W are valid, reliable and responsive measures in child and adolescent populations aged 8 - 16 years with anterior knee pain. These PROMs (Patient Reported Outcome Measures) can now be used in anterior knee pain conditions 8 years and above.

*Level of Evidence:* II Prospective pre- and post-test methodological study

**Contributions of the paper:**

- The PROMs-AKP, LEFS, VAS-U and VAS-W can be used from 8 years old to adulthood in anterior knee pain.
- This study provides evidence showing the above PROMs are valid and reliable in the child and adolescent population with anterior knee pain.
- These PROMs now have the potential to measure and monitor performance in children with anterior knee pain as has been done in adults.

## Introduction

Paediatric patients with symptoms of ‘Anterior knee pain’ commonly present to emergency and orthopaedic outpatients departments at Australian public tertiary/quaternary hospitals. Anterior knee pain in the paediatric patient is associated with high reoccurrence rates and treatment results are not always positive (Rathleff et al, 2019). Long term consequences of anterior knee pain include pain, reducing or ceasing sport participation, increased risk of adiposity, affecting career choices and health behaviour can be impacted (Rathleff et al, 2019). Acute knee injuries have been shown to predispose an individual to arthritis in later life (Maniar, Verhagen, Bryant & Opar, 2022).

Condition specific patient reported outcome measures (PROMs) are widely administered to adult patients who present with AKP during their clinical management journey. Commonly used and validated PROMs in the adult population for AKP include the Anterior Knee Pain Scale (AKPS), the Lower Extremity Functional Scale (LEFS) and the Visual Analog Scale Usual and Worst (VAS-U, VAS-W) (Crossley, Bennell, Cowan & Green, 2004; Watson, Propps, Ratner et al. 2005: Bennell, Bartam, Crossley & Green, 2000; Kujala, Jaakkola, Koskinen, et al. 1993; Crossley, Cowan, McConnell & Bennell 2005; Binkley, Stratford, Lott & Riddle 1999; Chesworth, Culham, Tata & Peat, 1989). AKP specific PROMs are less frequently used in the paediatric population as their clinimetric properties have not been established for this age group. Unfortunately, currently validated paediatric knee outcome measures such as the PEDi -IDKC (Pedi International Kee Documentation Committee) and KOOS (Knee Injury and Osteoarthritis Outcome Score) do not focus on AKP and therefore they are not sensitive change. For this reason, Myer et al. (2016) advocated that future research be conducted to determine whether adult AKP PROMs could be administered in the child and adolescent population. To the authors’ knowledge, no previous studies have evaluated the use of adult AKP PROMs in children under 14 years old who present with AKP. Therefore, the purpose of this study was to determine whether PROMs commonly used for adults experiencing AKP demonstrate acceptable clinimetric properties when utilized with children and adolescents. We hypothesized that the AKPS, LEFS, VAS-U and VAS-W would demonstrate acceptable clinimetric properties (i.e., reliability, validity, and responsiveness) in patients with knee pain that are aged 8 and above. The findings of this study will inform the PROMs that can be confidently used in daily practice for clinicians in tertiary paediatric hospitals.

## Method

### Study design

This prospective study involved analysis of PROMs collected from children attending the Queensland Children’s Hospital (QCH) and the Mater Hospital Brisbane physiotherapy services 2020-2021 period.

### Participants

Participants included children and adolescents: (1) aged 8-16 years, (2) referred to the QCH or the Mater Hospital Brisbane for physiotherapy management, (3) with anterior knee pain, pain as main driver. Children were excluded if they had (1) previous knee surgery (2) Clinical signs of knee pathology other than AKP (any instability as primary driver, internal derangement or bony pathology) (3) physiotherapy for AKP in previous 2 months 4. Two or more subluxations or dislocations. Data was collected using a prospective sample. Institutional ethics approval was gained from the Mater Health Service Human Research Ethics Committee and each patients / guardian provided informed written consent. Furthermore, the participants and the parent/guardian were encouraged to discuss if they would like the parent/guardian to be present at data collection.

### Measures and procedures

The participants (child or adolescent) completed the AKPS, LEFS, VAS-U and VAS-W in random order. These questionnaires were completed (1) at the beginning of the initial physiotherapy session, (2) at home 2-3 days later and returned at the next physiotherapy session, and (3) at the final physiotherapy visit, as per standard care along with the GRC (11 point scale).

#### Instruments

The Visual Analogue Scale (VAS) is a patient perceived pain measure first described by Scott & Huskisson in 1976 (13). It has minimum and maximum defined limits of self-perceived pain (D’Eon, Harris, & Ellis, 2004). Chesworth et al. (1989) validated the outcome measure for patellofemoral syndrome.

The Anterior Knee Pain Scale (AKPS) is a self-reported measure that was first introduced for adult females with knee pain and demonstrated construct and content validity (Kujala, Jaakkola, Koskinen, et al. 1993). The AKPS has concurrent validity, test-retest reliability, responsiveness in adults (Watson, Propps, Ratner, et al. 2005; Crossley, Cowan, McConnell & Bennell, 2005) and a strong correlation between the criterion measure of change (Smith, Selfe, Thacker, et al. 2018).

Binkley, Stratford, Lott & Riddle (1999) launched the Lower Extremity Functional Scale (LEFS) as a questionnaire evaluating a person’s ability to perform functional tasks. The LEFS showed test-retest reliability and construct validity (Bennell, Bartam, Crossley & Green, 2000) and test-retest reliability and responsiveness (Watson, Propps, Ratner, et al. 2005). The LEFS demonstrated high test-retest reliability and a significant correlation between the criterion measure of change with an ICC (intraclass correlation confidence) of 0.98 (Watson, Propps, Ratner, et al. 2005).

Global Rating of Change (GRC) assesses change over time for patients with pain (Watson, Propps, Ratner, et al. 2005). The form asks the therapist and the patient basic questions to help predict the degree of improvement or non-improvement in patients, with 7-11 point scale having the best test re-test reliability, patient preference and adequate discriminative ability (Kamper, Maher & Mackay, 2009).

### Statistical Analyses

#### Sample Size and statistical power

A sample size was estimated based on the test-retest reliability, with 49 patients required to achieve 80% power with a significance level of 0.05 for an expected reliability (intraclass correlation coefficient (ICC_3,1_)) of 0.80, when the minimum expected reliability (ICC_3,1_) is 0.60, and with 2 observations per subject (Walter, Eliasziw & Donner, 1998).

#### Statistical methods including data analysis

1. Patients’ characteristics were summarised by frequency and percentage for categorical variables and by mean and standard deviation (SD) or median (IQR) if non-normally distributed, for continuous characteristics, Statisical analyses were performed in Stata version 15 (Stata Corp, 2017).

### Global Rating of Change (GRC) Scores – 11 point scale

The agreement of the final GRC measure (around three months after initial), dichotomised to incomplete recovery had a GRC<5 and complete recovery GRC=5, was assessed between each combination of patient, therapist and parent/guardian using Cohen’s kappa. For each patient, the combined GRC score was assessed as a complete recovery only with agreement of all three raters (patient, parent and therapist).

### Reliability

Intraclass correlation coefficient (ICC), calculated based on single measures using two-way mixed-effects models with absolute agreement, and paired *t* tests were implemented to calculate the test-retest reliability using the initial and follow-up (2-3 days later) questionnaire score for the questionnaires. The *standard error (SE) of measurement (SEM)* was calculated to measure the degree of error in absolute terms and to express it in a scale of each outcome measure [SEM = SD_base_ x √2(1-R)]. The minimal detectable change was calculated using test-retest reliability coefficient to estimate the SEM. [MDC_95_ = 1.96 x SEM]. Reliability according to ICC show values of moderate = 0.5 and 0.75 good = 0.75 and 0.9 and excellent =0.9^25^.

### Validity

Trends in the final scores, change scores and relative change scores against baseline levels were compared graphically to determine whether analyses for validity should be adjusted for baseline levels.

The convergent (construct) validity, or whether the measures correlated with an external indicator of complete recovery as determined by combined GRC categories (incomplete recovery, complete recovery), were assessed using analysis of covariance (ANCOVA), with adjustment for baseline scores.

### Responsiveness

Responsiveness, or the ability to detect important change was assessed using an ANCOVA, adjusted for baseline scores between GRC categories.

Change scores were defined as the final score minus the baseline score. Receiver-operator characteristic (ROC) curve with the Youden index were also used to assess responsiveness and the change score cut-off predictive of complete improvement as per the GRC. The area under the curve (AUC), sensitivity and specificity to detect complete improvement at this change score cut-off were determined, and this cut-off was used to inform the minimal clinically important difference. The minimal clinically important difference was also assessed using the median change score (IQR) for each of the combined GRC categories (incomplete recovery, complete recovery), with associations assessed using the Mann-Whitney U test.

## Results

### Participant characteristics

Consent was gained from all the 50 participants included in analysis, 62% (n=31) were female, 60% (n=30) over the age of 13 years and most patients (n=29, 58%) experienced ‘anterior knee pain’ as their primary condition which includes SLJ, OGS, and PFPS, 42% had patella subluxation or first time patella dislocation, 0% had patella tendinopathy in this cohort (Table 1).

**Table 1.** Patient Characteristics.

|  | Total | Complete Improvement (GRC=5) |  | p-value |
| --- | --- | --- | --- | --- |
|  | n=50 | No<br>n=31 | Yes<br>n=19 |  |
| Gender, n (%) |  |  |  | 0.77 |
| Male | 19 (38%) | 11 (35%) | 8 (42%) |  |
| Female | 31 (62%) | 20 (65%) | 11 (58%) |  |
| Age Group, n (%) |  |  |  | 1.00 |
| 8 to 12yrs | 20 (40%) | 12 (39%) | 8 (42%) |  |
| 13yr + | 30 (60%) | 19 (61%) | 11 (58%) |  |
| Condition, n(%) |  |  |  | 0.79 |
| Patella sublux/ dislocations | 10 (20%) | 7 (23%) | 3 (16%) |  |
| Anterior knee pain | 29 (58%) | 18 (58%) | 11 (58%) |  |
| Patella dislocations | 11 (22%) | 6 (19%) | 5 (26%) |  |
| Sport Group, n (%) |  |  |  | 1.00 |
| Nil | 19 (38%) | 12 (39%) | 7 (37%) |  |
| School PE | 31 (62%) | 19 (61%) | 12 (63%) |  |

Note: ≥ greater than or equal to, IQR = median change score

Questionnaire score summary of the AKPS, LEFS, VAS-U and VAS-W results can be seen in Table 2.

**Table 2.** Questionnaire score summary.

| Measure | Initial | 2-3 days after initial | 3 months |
| --- | --- | --- | --- |
|  | median (IQR) | median (IQR) | median (IQR) |
| VAS-U (0-10) | 7.0 (5.0-7.0) | 6.0 (5.0-7.0) | 0.0 (0.0-1.0) |
| VAS-W (0-10) | 8.0 (7.0-9.0) | 8.0 (7.0-8.0) | 1.0 (0.0-2.0) |
| AKPS (0-100) | 42.5 (27.0-56.0) | 42.0 (29.0-57.0) | 100.0 (92.0-100.0) |
| LEFS (0-80) | 29.0 (16.0-40.0) | 29.0 (17.0-41.0) | 80.0 (78.0-80.0) |

Note: VAS-U/ VAS-W Visual Analogue Scale Usual/ Worst, AKPS Anterior Knee Pain Scale, LEFS Lower Extremity Functional Scale, IQR = median change score

Ideally studies compare improvement versus no change or worsening but all patients improved within the study duration, so data did not have a wide range of GRC scores. The median change score summary at 3 months was 5.0 for the patient, 4.0 for the therapist and 4.5 for the parent. Due to this this study looked at complete improvement on the GRC versus incomplete improvement on the GRC. Agreement between patient and therapist, and patient and parent were moderate (k=0.47, k=0.48, respectively). Agreement between parent and therapist were highly correlated k=0.80), with only five parents disagreeing with therapists and considering their child completely healed. The largest discrepancies in agreement were cases of patients considering themselves completely healed. Nineteen patients were considered completely improved using the combined GRC scores (Table 3).

**Table 3.**
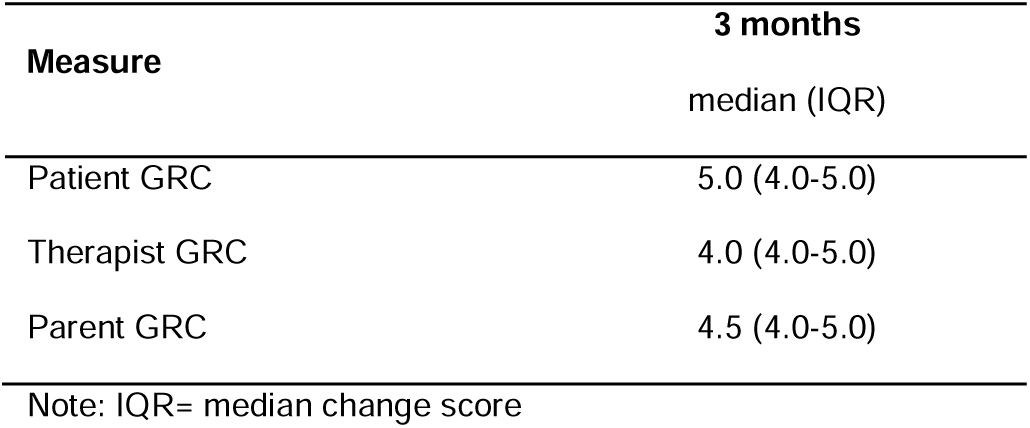
Global Rating of Change (GRC) Questionnaire score summary at 3 months.

| Measure | 3 months |
| --- | --- |
|  | median (IQR) |
| Patient GRC | 5.0 (4.0-5.0) |
| Therapist GRC | 4.0 (4.0-5.0) |
| Parent GRC | 4.5 (4.0-5.0) |

Note: IQR= median change score

### Test-retest reliability

The test-retest summary and minimal detectable change results are shown in Table 5. ICC value for measures were 0.86 for VAS-U indicating good reliability; 0.70 for the VAS-W, indicating moderate reliability; and 0.98 for both AKPS and LEFS indicating excellent reliability. There was a significant mean difference score between the first two surveys for VAS-U, VAS-W and LEFS, but not for the AKPS. The magnitude of the difference was smaller than the SEM.

The minimal detectable change based on the SEM required to detect a change greater than the measurement error 95% of the time in this population was 1.7 units for VAS-U, 2.2 units for VAS-W, 9.0 units for AKPS and 6.5 units for LEFS. The percentage change in terms of the total scores were VAS-U 27.5%, VAS-W 28.5%, AKPS 21.6% and LEFS 21.4%.

### Validity

Plots of the final (3-month) change score for each of the measures are shown in Figure 1. For all measures, a large proportion of patients reach the maximum score at 3 months (floor and ceiling effects: VAS-U/VAS-W = 0, AKPS = 100, LEFS = 80) across the full range of baseline measure values; VAS-U 62% (n=31), VAS-W 44% (n=22), AKPS 52% (n=26), LEFS 56% (n=28).

**Figure 1.**
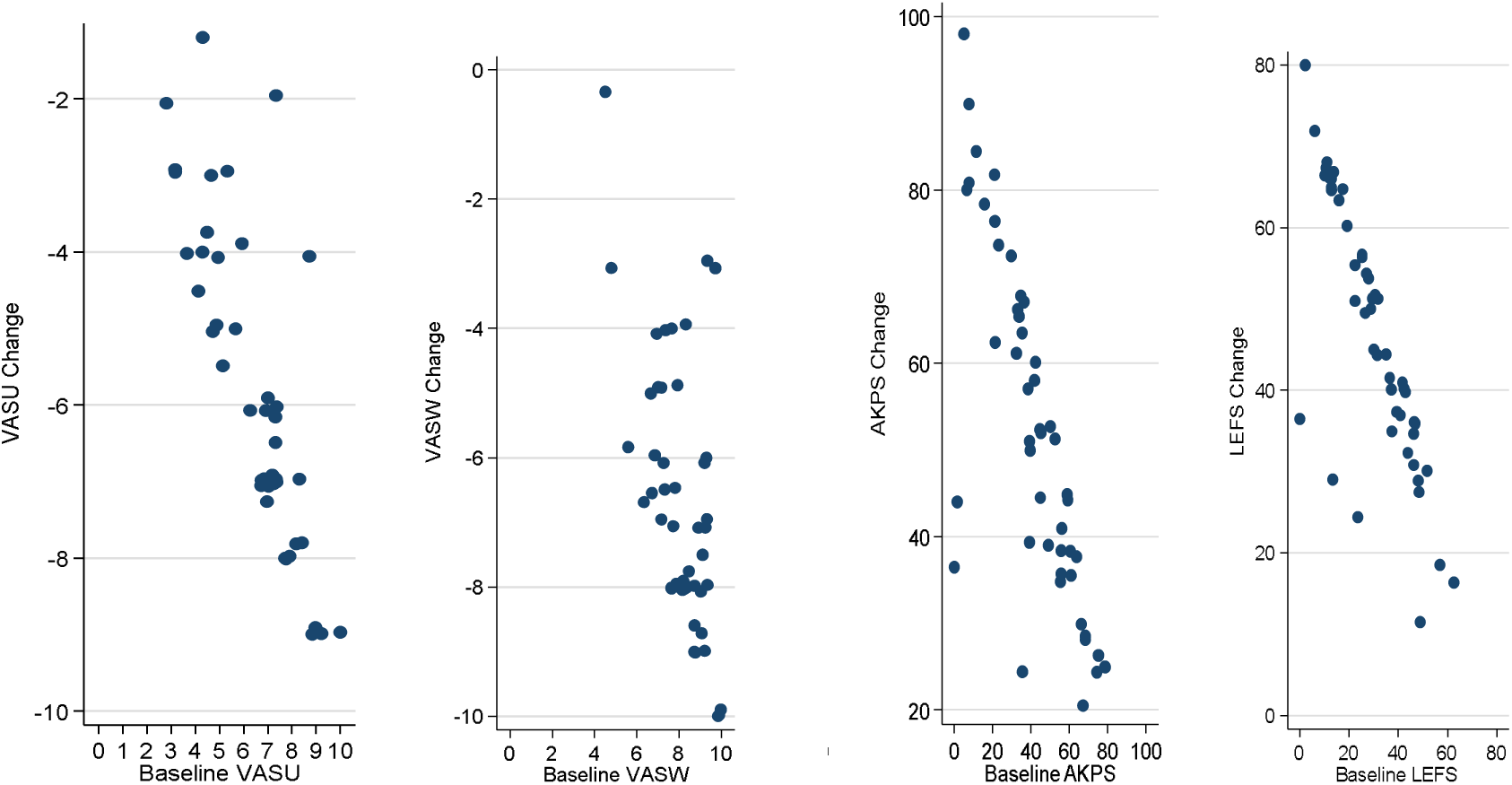
Visual Analogue Scale Usual (VAS-U)/ Worst (VAS-W), Anterior Knee Pain Scale (AKPS), Lower Extremity Functional Scale (LEFS) improvement compared to baseline.

The change scores for each of the measures were linearly associated with the baseline scores, with larger change scores observed for patients with worse baseline scores regardless of measure. VAS-U and VAS-W scores moving closer to zero indicate improvement (negative change score), while AKPS and LEFS scores moving closer to 100 and 80, respectively, (positive change score) indicated improvement.

The ANCOVA compared the baseline score adjusted mean change scores of the measures between participants who exhibited complete improvement, through the combined GRC at 3 months, with those who did not (Table 4). In the combined GRC, 38% (n=19) of patients exhibited complete improvement (Table 6).

**Table 4.** Patient, Parent, Therapist Global Rating of Change (GRC) complete improvement agreement.

| Rater 1 | Rater 2 | Raters Agree - Yes | Raters Agree - No | Rater 1 - Yes<br>Rater 2 - No | Rater 1 - No<br>Rater 2 - Yes | Agreement | Kappa (95% CI) |
| --- | --- | --- | --- | --- | --- | --- | --- |
| Patient | Therapist | 19 | 17 | 13 | 1 | 72.0% | 0.47 (0.26-0.68) |
| Parent | Patient | 22 | 15 | 3 | 10 | 74.0% | 0.48 (0.25-0.71) |
| Therapist | Parent | 20 | 25 | 0 | 5 | 90.0% | 0.80 (0.64-0.98) |

Note: CI Confidence Interval, Agreement= same GRC score to each other

**Table 5.** Summary of test-retest reliability and minimal detectable change.

| Measure | Survey 1 | Survey 2 | Paired t test |  |  | ICC (95% CI) | SEM | MDC |
| --- | --- | --- | --- | --- | --- | --- | --- | --- |
|  | Median (IQR) | Median (IQR) | Mean difference (95% CI) | T(49) | P-value |  |  |  |
| VAS-U | 7.0 (5.0-7.0) | 6.0 (5.0-7.0) | 0.5 (0.3 - 0.7) | 4.9 | <0.001 | 0.86 (0.61 - 0.94) | 0.62 | 1.7 |
| VAS-W | 8.0 (7.0-9.0) | 8.0 (7.0-8.0) | 0.6 (0.4 - 0.9) | 4.5 | <0.001 | 0.70 (0.39 - 0.85) | 0.80 | 2.2 |
| AKPS | 42.5 (27.0-56.0) | 42.0 (29.0-57.0) | 0.1 (-1.2 - 1.4) | 0.1 | 0.90 | 0.98 (0.96 - 0.99) | 3.25 | 9.0 |
| LEFS | 29.0 (16.0-40.0) | 29.0 (17.0-41.0) | -1.2 (2.1 - -0.3) | -2.7 | 0.009 | 0.98 (0.96 - 0.99) | 2.34 | 6.5 |

**Table 6.**
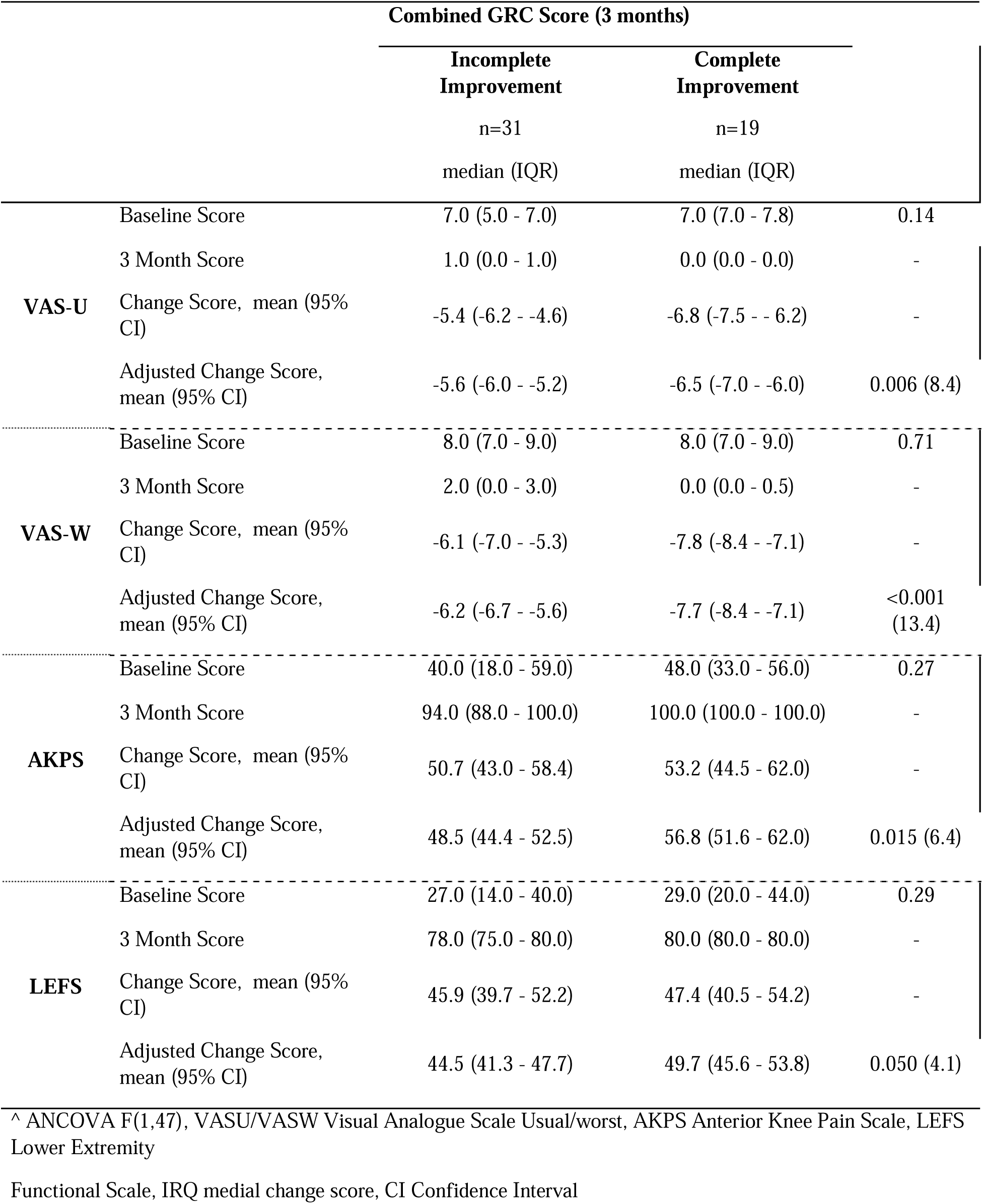
Changes in outcome measures corresponding to complete or incomplete improvement in the Global rating of change.

There was very strong evidence of a difference in the adjusted mean change score for both the VAS-U (p=0.006) and the VAS-W (p<0.001) for improvement groups, with a larger mean change exhibited in the complete improvement group. There was moderate evidence of a difference in the adjusted mean change score for the AKPS measure (p=0.015), and some evidence for the LEFS measure (p=0.050), with larger mean adjusted change scores but the confidence intervals largely overlapped.

Significantly larger adjusted mean scores were noted in the complete group compared to the incomplete improvement group, suggesting convergent validity of the measures.

There was no difference in the baseline scores between the GRC improvement groups as evidenced in Table 6.

### Responsiveness

Responsiveness was also evaluated through the ANCOVA results (Table 6). The median change scores are seen on Table 6. The significantly larger adjusted mean scores in the complete recovery group compared to the incomplete recovery group, suggest these measures are responsive to distinguish complete recovery from incomplete recovery when adjusted for patient baseline scores. The VAS-U mean scores at 3 months were lower in both the incomplete improvement (5.6 units) and complete improvement (6.5 units) groups after adjusting for baseline scores. The VAS-W mean scores at 3 months were lower in both the incomplete improvement (6.2 units) and complete improvement (7.7 units) groups after adjusting for baseline scores. The AKPS mean scores at 3 months were higher in both the incomplete improvement (48.5 units) and complete improvement (56.8 units) groups after adjusting for baseline scores. The LEFS mean scores at 3 months were higher in both the incomplete improvement (44.5 units) and complete improvement (49.7 units) groups after adjusting for baseline scores.

Table 7 summarises the ROC analysis on the measure change scores and complete improvement on the combined GRC; including the area under the curve (AUC), sensitivity and specificity to detect complete improvement, and the change score cut-off required to detect complete change.

**Table 7.** Receiver operating characteristics (ROC) for all measures.

| Measure | AUC (95% CI) | Sensitivity | Specificity | Change Score Cut-off | Percentage Change Score |
| --- | --- | --- | --- | --- | --- |
| VAS-U | 0.71 (0.57 - 0.85) | 0.79 | 0.65 | -6.2 | 62% |
| VAS-W | 0.74 (0.61 - 0.88) | 0.95 | 0.52 | -6.0 | 60% |
| AKPS | 0.56 (0.40 - 0.72) | 0.74 | 0.45 | 41 | 41% |
| LEFS | 0.52 (0.35 - 0.68) | 0.89 | 0.26 | 32 | 40% |

Note: AUC Area Under the Curve, CI Confidence Interval, VAS-U VAS-W Visual Analogue Scale Usual/ Worst, AKPS Anterior Knee Pain Scale, LEFS Lower Extremity Functional Scale

The measure best able to distinguish between complete and incomplete improvement on the combined GRC was the VAS-W with an AUC of 0.74, demonstrating some discriminatory power. This measure had very high sensitivity (95%) but low specificity (52%), indicating 48% of those above the cut-off will be false positive results. The LEFS was least able to distinguish between complete and incomplete improvement with an AUC of 0.52, demonstrating poor discriminatory power; with 74% classed as improved by the change score cut-off considered false positives. The percentage change score required to detect complete improvement were calculated for VAS-U (62%) and VAS-W (60%) AKPS (41%) and LEFS (40%). This suggests that only VAS-U and VAS-W scores are responsive to detect complete improvement in patients, without considering baseline scores.

There was no evidence of associations with the adjusted mean change score of the measures by age, gender or patient condition. Associations of lower baseline AKPS scores (greater pain) in patients with ‘patella dislocations’ than those with ‘anterior knee pain’ and patella subluxation/dislocations, were however found see Table 8 and 9.

**Table 8.** Associations with outcome measures and patient condition.

| Measure |  | Patella subluxation/<br>dislocations | Anterior knee pain | Patella dislocations | p-value (F<sup>^</sup>) |
| --- | --- | --- | --- | --- | --- |
|  |  | n=10 | n=29 | n=11 |  |
|  |  | mean (95% CI) | mean (95% CI) | mean (95% CI) |  |
| VAS-U | Baseline, median (IQR) | 7.0 (4.2 - 7.0) | 7.0 (6.0 - 7.0) | 7.0 (5.0 - 9.0) | 0.42 |
|  | Change | -5.1 (-6.8 - -3.3) | -6.1 (-6.7 - -5.4) | -6.3 (-7.8 - -4.9) | - |
|  | Adjusted change | -5.6 (-6.3 - -4.8) | -6.0 (-6.5 - -5.6) | -5.9 (-6.6 - -5.2) | 0.57 (0.6) |
| VAS-W | Baseline, median (IQR) | 8.5 (7.0 - 9.0) | 8.0 (7.0 - 9.0) | 8.6 (8.0 - 9.0) | 0.38 |
|  | Change | -6.4 (-8.4 - -4.4) | -6.7 (-7.3 - -6.0) | -7.3 (-8.8 - -5.9) | - |
|  | Adjusted change | -6.4 (-7.5 - -5.3) | -6.8 (-7.5 - -6.2) | -6.9 (-8.0 - -5.9) | 0.76 (0.3) |
| AKPS | Baseline, median (IQR) | 32.5<br>(11.0 - 54.0) | 49.0<br>(40.0 - 64.0) | 19.0<br>(8.0 - 40.0) | 0.002 |
|  | Change | 58.8<br>(39.5 - 78.1) | 45.2<br>(39.7 - 50.8) | 62.0<br>(48.7 - 75.3) | - |
|  | Adjusted change | 51.9<br>(44.2 - 59.7) | 52.6<br>(47.8 - 57.4) | 48.8<br>(40.9 - 56.7) | 0.73 (0.3) |
| LEFS | Baseline, median (IQR) | 22.5<br>(14.0 - 44.0) | 34.0<br>(26.0 - 40.0) | 16.0<br>(10.0 - 44.0) | 0.096 |
|  | Change | 48.3<br>(32.7 - 63.9) | 44.4<br>(39.5 - 49.3) | 50.3<br>(38.5 - 62.1) | - |
|  | Adjusted change | 44.6<br>(38.7 - 20.4) | 48.2<br>(44.7 - 51.7) | 43.7<br>(38.0 - 49.4) | 0.34 (1.1) |

Note: ^ANCOVA F(2,46); IQR median change score, CI Confidence Interval, VAS-U VAS-W Visual Analogue Scale Usual/ Worst, AKPS Anterior Knee Pain Scale, LEFS Lower Extremity Functional Scale

**Table 9.** Associations with outcome measures and age.

|  |  | 8 to 12yrs | 13yr + | p-value (F<sup>^</sup>) |
| --- | --- | --- | --- | --- |
| Measure |  | n=20<br>mean (95% CI) | n=30<br>mean (95% CI) |  |
| VASU | Baseline median (IQR) | 7.0 (7.0 - 7.0) | 7.0 (5.0 - 7.8) | 0.22 |
|  | Change | -6.5 (-7.2 - -5.9) | -5.5 (-6.3 - -4.7) |  |
|  | Adjusted Change | -6.2 (-6.7 - -5.6) | -5.8 (-6.2 - -5.4) | 0.27 (1.3) |
| VASW | Baseline median (IQR) | 8.0 (7.5 - 8.9) | 8.0 (7.0 - 9.0) | 0.94 |
|  | Change | -7.2 (-8 - -6.5) | -6.4 (-7.3 - -5.6) |  |
|  | Adjusted Change | -7.2 (-7.9 - -6.4) | -6.5 (-7.1 - -5.9) | 0.14 (2.2) |
| AKPS | Baseline median (IQR) | 41.0 (26.0 - 63.0) | 45.0 (27.0 - 55.0) | 0.71 |
|  | Change | 51.6 (42.9 - 60.3) | 51.7 (43.8 - 59.5) |  |
|  | Adjusted Change | 52.4 (47.0 - 57.7) | 51.1 (46.7 - 55.5) | 0.72 (0.1) |
| LEFS | Baseline median (IQR) | 31.5 (18.0 - 44.0) | 26.5 (15.0 - 40.0) | 0.47 |
|  | Change | 45 (38.1 - 51.9) | 47.5 (41.2 - 53.7) |  |
|  | Adjusted Change | 46.9 (42.7 - 51.1) | 46.2 (42.8 - 49.6) | 0.8 (0.7) |

Note: <sup>^</sup>ANCOVA F(1,47); IQR median change score, CI Confidence Interval, VAS-U VAS-W Visual Analogue Scale Usual/ Worst, AKPS Anterior Knee Pain Scale, LEFS Lower Extremity Functional Scale

## Discussion

The purpose of this study was to determine whether PROMs commonly used for adults experiencing AKP demonstrate acceptable clinimetric properties when utilized with children and adolescents. In agreement with our hypothesis, the outcome measures VAS-U, VAS-W, AKPS and LEFS with their respective psychometric properties were valid and reliable outcome measures for anterior knee pain presentations in 8-16-year-olds and could be used for clinical and research purposes.

The VAS-U and VAS-W questionnaires demonstrated high construct validity and the AKPS and LEFS questionnaires demonstrated moderate construct validity and internal consistency in AKP patients aged from 8-16 years. There were ceiling and floor effects observed for all questionnaires with most of the cohort recording complete recovery from the start to the end of their intervention (see Table 2). These changes indicate that the instruments were responsive to changes in function and were able to measure the magnitude of change (Irrgang, Anderson, Boland, et al, 2001). However, the observed floor and ceiling effect at the final timepoint may indicate that the questionnaires were not sensitive enough to demonstrate meaningful clinical change in this population group.

All PROMs were reliable in assessing AKP in this population with excellent reliability in AKPS and LEFS, good reliability in VAS-U and moderate reliability in VAS-W. This indicates these PROMs can be reliably used to assess change over time with or without treatment. Given, similar reliability results of these PROMs have been shown in adult populations with patellofemoral pain conditions (Bennell, 2000, Watson, 2005). These PROMs could be used to assess change of patellofemoral conditions over the lifespan, giving insight into the condition from youth to adulthood.

At first glance, the significant mean difference score between initial questionnaire scores and questionnaire scores 2-3 days later for VAS-U, VAS-W and LEFS might suggest testing variability, however the change for all PROMs were in the direction of less pain for all measures, suggesting this could be due to minor improvements during this short time of 2-3 days. Thus, the observed differences were likely systematic difference rather than variability in the PROMs and should be considered when interpreting these PROMs in younger patients who may experience faster recovery times compared to adults. The VAS-U and VAS-W were responsive to detect complete improvement using change scores. With baseline adjustment the AKPS and LEFS were found to be responsive.

Kujula, Jaakkola, Koskinen, et al. (1993) designed the AKPS in 1993 and focused their study on subjects with anterior knee pain conditions, patella subluxations or patella dislocations and controls in the adult population. Myer et al. (2016) went to show that the AKPS was valid and reliable in young female athletes from 11 to 18 years of age. In this study the patellofemoral instability group and the anterior knee pain populations were combined. The results did not show any differences in the groups in this study with the adjusted change scores for any of the outcome measures (Table 6). This indicates that there was no difference in the outcome measure results seen in patients with Patella instability/1^st^ time patella dislocators versus those with anterior knee pain alone in the paediatric population studied.

There are several considerations that should be taken into account when interpreting the findings of this study. Firstly, patient improvement over the study duration only allowed assessment of complete improvement instead of general/partial improvement. Further research with final assessment done earlier may be needed to assess responsiveness of PROMs to detect general improvement without resolution of symptoms. Further to this in the early stages of this research, investigators realized that parents had to explain some technical terms in the questionnaires like “flexion”, “flexion deficiency” and “atrophy” in the AKPS questionnaire. Myer et al. (2016), recommended the AKPS terminology may require amendment in the adolescent population, as it reduces independent completion in this age group. Vitale, Levy, Johnson, Gelijns et al. (2001), evaluated quality of life assessments in the adolescent population and concluded specifically designed tools are required to reflect the unique needs of this population as they are fundamentally different to adults and their health status. Hence, development of the paediatric versions of the IKDC (International Knee Documentation Committee Subjective Knee Evaluation Form) (Irrgang, Anderson, Boland et al, 2001) and KOOS -Child (Knee Injury and Osteoarthritis Outcome Score-Child) (Ortqvist, Roos, Brostrom, Janarv, Iversen, 2012) have allowed easier understanding of the paediatric population (van der Velden, van der Steen, Leenders, van Douveren et al, 2019). Similar modifications to the AKPS and the LEFS may be warranted to allow easier understanding in the paediatric population. Further research could also include using reduced questionnaires such as the 6-item AKPS developed by Myer et al. (2016) for the adolescent population which may improve compliance to completing the questionnaires independently or adapting the existing questionnaires to improve readability and understanding in the child and adolescent population. This may improve responsiveness of questionnaires to allow more accurate clinical application.

## Conclusion

Like adults, the AKPS, LEFS, VAS-U, and VAS-W are valid, reliable and responsive measures in the child and adolescent population aged 8 - 16 years with anterior knee pain. Clinicians and researchers can be confident these outcome measures effectively evaluate clinical efficacy in the management of 8-16-year-olds with anterior knee pain. This now enables clinicians to be able to use these PROMs in anterior knee pain conditions across the child, adolescent and adult populations.

## Implications of Physiotherapy Practice

Provide valid and reliable outcome measures that can be used from child to adult in the anterior knee pain population.

Allow monitoring and measuring change over time for clients with anterior knee pain for clinicians and researchers.

Monitor and measure symptoms or activities that are known to correlate with anterior knee pain across child to adult populations

## Data Availability

All data produced in the present study are available upon reasonable request to the authors

